# Time trends of mental health indicators in Germany’s adult population before and during the COVID-19 pandemic

**DOI:** 10.1101/2022.10.09.22280826

**Authors:** Elvira Mauz, Lena Walther, Stephan Junker, Christina Kersjes, Stefan Damerow, Sophie Eicher, Heike Hölling, Stephan Müters, Diana Peitz, Susanne Schnitzer, Julia Thom

## Abstract

**Background:** Times of crisis such as the COVID-19 pandemic are expected to compromise mental health. Despite a large number of studies, evidence on the development of mental health in general populations during the pandemic is inconclusive. One reason may be that representative data spanning the whole pandemic and allowing for comparisons to pre-pandemic data are scarce.

**Methods:** We analyzed representative data from telephone surveys of Germany’s adults: “German Health Update (GEDA)” and “COVID-19 vaccination rate monitoring in Germany (COVIMO)”. Three indicators of mental health were observed in approximately 1,000 and later 3,000 randomly sampled participants monthly until June 2022: symptoms of depression (observed since April 2019 using the PHQ-2), symptoms of anxiety (observed since March 2021 using the GAD-2) and self-rated mental health (observed since March 2021 using a single item). We produced time series graphs including estimated three-month moving means and proportions of individuals with a positive screen (PHQ/GAD-2 score ≥ 3) and of those reporting very good/excellent mental health, as well as smoothing curves. We also compared time periods between years. Analyses were stratified by sex, age, and level of education.

**Results:** While mean depressive symptom scores declined from the first wave of the pandemic to summer 2020, they increased from October 2020 and remained consistently elevated throughout 2021 with another increase between 2021 and 2022. Correspondingly, the proportion of positive screens first decreased from 11.1 % in spring/summer 2019 to 9.3 % in the same period in 2020 and then rose to 13.1 % in 2021 and to 16.9 % in 2022. While depressive symptoms increased in all subgroups at different times, developments among women, the youngest and eldest adults, and the high level of education group stand out. Furthermore, symptoms of anxiety increased while self-rated mental health decreased between 2021 and 2022.

**Conclusions:** Elevated symptom levels and reduced self-rated mental health at the end of our observation period in June 2022 call for further continuous mental health surveillance. Mental healthcare needs of the population should be monitored closely. Findings should serve to inform policymakers and clinicians of ongoing dynamics to guide health promotion, prevention, and care.

## 1 Introduction

### The Covid-19 pandemic poses a serious threat to mental health

Shortly after the World Health Organization declared the SARS-COV-2 outbreak a global pandemic on March 11, 2020 (1), alarms were sounded over a potential concomitant mental health crisis (2-4). A secondary pandemic in the form of a “tsunami of mental disorders” was expected, for example by the British Psychiatric Association (5). These assumptions were based on empirical evidence of population-wide increases in mental health risks associated with previous infectious outbreaks such as Ebola, influenza, and SARS (6-8), natural disasters (9), and economic crises (10, 11). Stressors accompanying infectious outbreaks include the experience of uncertainty and anxiety, threats or damage to physical health and potentially traumatic experiences such as the loss of loved ones. In addition to effects of the disease itself, nonpharmaceutical interventions (NPIs) to mitigate the spread of infections are discussed as contributing to mental health deterioration. As NPI-associated risk factors in the COVID-19 pandemic, the literature highlights isolation and quarantine (12, 13), an increase in domestic violence (14), and a lack of social connectedness during contact restrictions (15). Moreover, NPIs may lead to the loss of protective factors for mental health such as social and recreational activities and access to healthcare (16). In addition to these individual-level factors, societal-level mental health risks such as economic strain resulting in increased unemployment and the risk of widening social inequality are likely to arise from the pandemic (16-18). Against this background, the COVID-19 pandemic is considered a multidimensional and now chronic stressor continuously putting the mental health of populations at risk (16, 19).

### Like most countries, Germany has been hit by multiple waves of rising COVID-19 incidence and mortality as well as NPIs in response, which might relate to mental health dynamics temporally

Taking various epidemiological, healthcare- and policy-related parameters into account, the course of the pandemic in Germany can retrospectively be divided into eight phases (see Figure 1) (20-24). After the first confirmed SARS-CoV-2 infection on 27^th^ January, 2020, a nationwide first wave of infections followed (March to May 2020). NPIs were put in place, resulting in an extensive lockdown which comprised contact and travel restrictions, cancellations of major events and closed childcare facilities, schools, and restaurants. A milder interim period of low case numbers referred to as a “summer plateau” (20, 23) followed from May to September 2020. From October 2020 to February 2021 a second, more severe wave unfolded, again met by several NPIs and the beginning of the vaccination campaign (24). After a brief period of declining case numbers, a third wave emerged from March to June 2021. Another short summer plateau in 2021 (June to July) was followed by a fourth wave of infections from August to December 2021 (21). With regard to COVID-19 incidence, the fourth wave was the most severe up to that point with a nearly 10-fold number of average cases per day compared to the first wave and a nonstop transition into wave five, which began in December 2021 (21-23) and was characterized by the highly contagious omicron variant with even higher infection rates (22, 25). Data on case numbers shows the largest peak to date in spring 2022 (26). NPIs, however, were largely lifted in most German federal states at the beginning of April 2022 (27).

**Figure 1:**
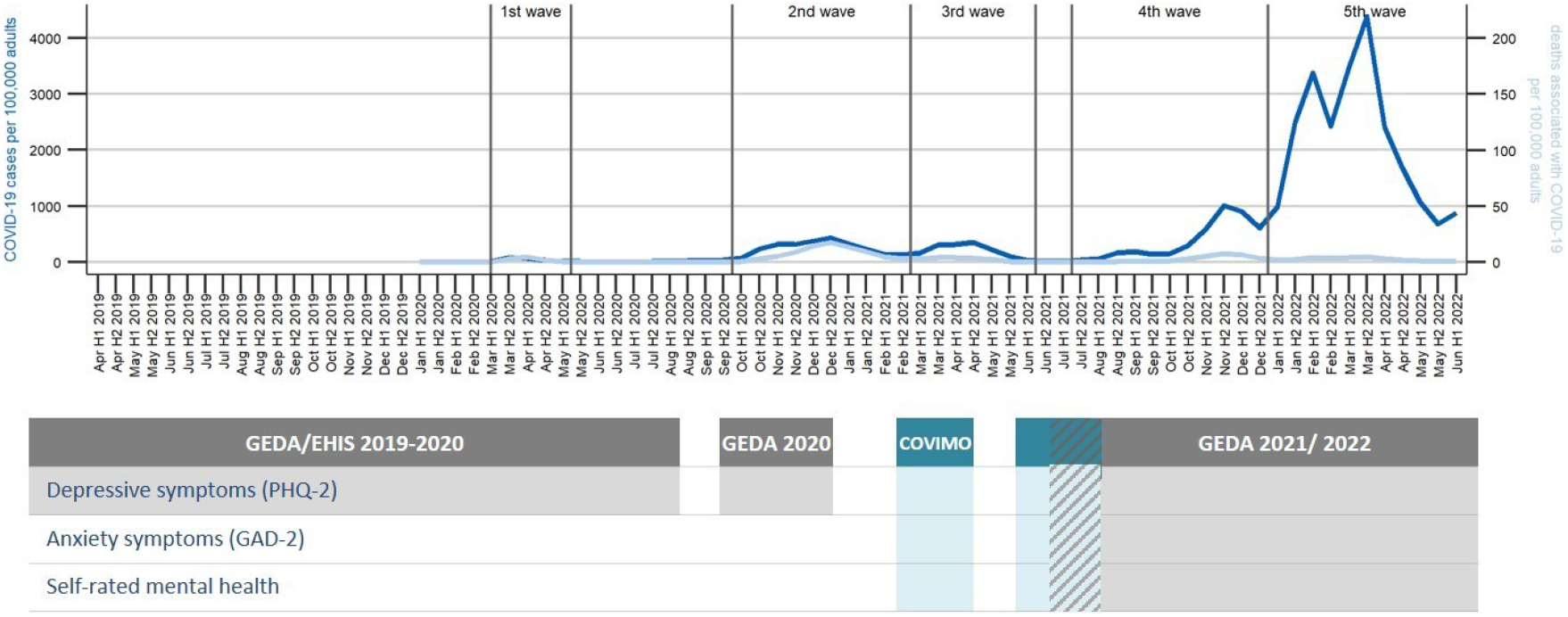
Data sources used in analyses over the course of the COVID-19 pandemic COVID-19 cases and associated deaths per 100,000 adults are calculated per first (H1) and second half (H2) of the month

Russia’s invasion of Ukraine on 24^th^ February, 2022, marks a further major event in this time and the beginning of another crisis on a global scale that might affect mental health dynamics.

### Despite the well-founded expectation of a mental health crisis, evidence on changes in mental health of adults during COVID-19 pandemic is (still) inconclusive

Turning first to international research, reviews point to a broad heterogeneity in current findings. While some reviews conducted early in the pandemic conclude that there was an increase in depressive and anxiety symptoms (28, 29), others found quick subsequent decreases or stable symptoms in general populations (30). A later review and meta-analysis reports no changes (31). One review summarizes a most likely ‘big picture’ (30) emerging from heterogeneous findings: symptom increases compared to pre-pandemic data during first lockdowns followed by declines as restrictions are eased, but not down to pre-pandemic levels (32). As findings accumulate, inconsistencies are growing, while the trajectory of manifest mental disorders remains an open question (33). A recent umbrella review based on 81 systematic reviews on global mental health trends during the pandemic evaluates the current state of research as follows: “Despite high volumes of reviews, the diversity of findings and dearth of longitudinal studies within reviews means clear links between COVID-19 and mental health are not available, although existing evidence indicates probable associations” (34, p. 2).

The existing literature from **Germany** prohibits clear conclusions as well. In a rapid review including 68 records published until mid-2021, we found study outcomes to be associated with the suitability of the data used for assessing changes in the general population reliably with regard to sampling methods and comparability of observation periods (35). While studies with particularly suitable research designs showed mixed results on the overall development of mental health in Germany, studies with more bias-prone designs predominantly reported deteriorating mental health. Importantly, two thirds of the reviewed studies are based on data collected during the first wave and the summer plateau of 2020, when COVID-19 incidence was comparatively low in Germany (35). The few studies that address the later course of the pandemic find an elevated frequency of depressive symptoms in the first months of 2021 (36, 37) or of depressive and anxiety symptoms into later 2020 (37, 38) compared to pre-pandemic data and an increase in mental distress (39) but a decrease in depressive and anxiety symptoms (37) in the second wave compared to the first wave. Further results from representative surveys spanning the whole pandemic period and allowing for comparisons to pre-pandemic baseline data are needed in order to adequately assess the mental health impact of the pandemic in the general population in Germany.

### Mental health developments in the pandemic may also vary by population subgroups. Although social inequalities in mental health already existed in non-pandemic times (40), there is evidence that they were aggravated by the COVID-19 pandemic (16, 19, 41, 42)

As expected, the widely observed **gender gap** resulting in mental health disadvantages for women compared to men was found to have worsened across a majority of studies (e.g. 41, 43, 44). A comprehensive meta-analysis on gender equality in the pandemic globally attributes this to unequally distributed pandemic-related risk factors such as an increase in domestic violence, increased childcare responsibilities, and financial losses (45). With regard to **age and life stages**, concerns have been raised for the mental wellbeing of the elderly due to the increased risk of severe COVID-19 disease progression (16, 19) and increased risk of loneliness and isolation due to a greater need for social distancing (19) in this group. However, increases in psychological distress and symptoms of mental illness have been predominantly reported for the youngest adults in particular (33, 41, 43, 46-49). One potential explanation is a larger impact of restrictions with a stronger disruptive effect in this transitional life phase (50). In Germany, women and younger adults have also been repeatedly observed to be more severely affected than men and other age groups (49, 51-54). By contrast, international empirical findings regarding **socioeconomic groups** and mental health during the COVID-19 pandemic have been inconsistent despite cumulative risks of individuals with a low socioeconomic status (SES). They face a greater risk of severe infection and death from COVID-19 (41, 42) and economic stressors such as financial insecurity, reduced working hours, and income or job loss (55, 56). Previous studies from different countries have shown mixed results, including low SES as a risk factor for depression and anxiety (57, 58), no association between SES and mental health (59, 60), and individuals with higher SES at a greater risk of worsening mental health in the pandemic (44, 51, 54, 61, 62). These discrepancies may be due to national contextual differences. Also, risk and resilience factors may change over time as circumstances change, calling for Germany-specific results on mental health by subgroup over the course of the pandemic.

### The dynamic nature of the COVID-19 pandemic and its high relevance to all areas of public health created specific informational needs with regard to the mental health of the population

Specifically, it calls for a public health surveillance approach (63) involving continuous observation and timely reporting of updated time trends as the basis for planning, implementing, and evaluating interventions to protect and promote the health of the population (64). Accordingly, public health authorities have set up ongoing population surveys in order to monitor mental health trends at high frequency and serve as an early warning system, for example in the US (65) and UK (66). In Germany, pre-pandemic mental health monitoring focused on the estimation of 12-month-prevalences of varying mental health indicators, based on health interview and health examination surveys conducted at perennial intervals (e.g. 67, 68-71). In 2019, the Federal Ministry of Health commissioned the Robert Koch Institute (RKI) to establish a national Mental Health Surveillance in order to provide systematic and continuous evidence on the mental health of the population. As a conceptual foundation, core indicators for public mental health were identified (72) and prioritized by national stakeholders (73), integrating international expertise (74). With the onset of COVID-19 pandemic, first indicators from the comprehensive set had already been implemented in the running field work of the survey “German Health Update (GEDA)” (70). As the pandemic progressed, further measures were added to GEDA as well as to “COVID-19 vaccination rate monitoring in Germany (COVIMO)” (75). This representative data from approximately 1,000 respondents per month, and, as of 2022, 3,000 per month for some indicators, makes tracking the development of several mental health indicators in the German population in high-frequency cross-sectional time series possible, addressing some of the above-mentioned research gaps.

**In the present study** we analyze month-by-month time series for symptoms of common mental disorders (depressive symptoms and anxiety symptoms) as well as an indicator of positive mental health (self-rated mental health) in order to address the following three research questions: (1) How did depressive symptoms develop between April 2019 and June 2022 in the adult population in Germany? (2) Did developments of depressive symptoms in the observation period differ by gender, age, and level of education? If so, did mental health differences between subgroups vary over time? (3) How did symptoms of anxiety disorders and self-rated mental health develop between March 2021 and June 2022? Importantly, we examine both mean depressive and anxiety symptom scores and proportions of the population screening positive for possible depressive or anxiety disorder. This allows us to distinguish between developments in symptom severity at the population level and changes in the percentage of the population with potentially clinically relevant symptom levels, both of which are important public health indicators (76).

## 2 Materials and Methods

### 2.1 Data

#### 2.1.1 Surveys

Figure 1 shows the phases of the COVID-19 pandemic in Germany and the data collection periods for the two surveys used and the three indicators analyzed in this study. At the start of the COVID-19 pandemic, the third survey wave of the European Health Interview Survey as part of the study “German Health Update” (GEDA 2019/2020-EHIS) for Germany (70) had been in the survey phase conducting telephone interviews since April 2019. This survey included the screening questionnaire PHQ-8 (77), which comprises its abbreviated version PHQ-2 (78), as a measure of depressive symptoms.

The survey was originally not designed for monthly reporting; however, slight adjustments of the sample weighting permitted first analyses of the development of various health indicators in the months preceding the pandemic as well as the first months of the pandemic (79). Given the new informational needs arising from the pandemic, the survey was continued until the beginning of January 2021. After the end of GEDA 2019/2020, short inventories assessing mental health indicators, including the PHQ-2 (78), the GAD-2 (80), and a self-rated mental health (SRMH) item (81) were integrated into a running population-based telephone survey from mid-March to mid-July 2021. This survey, the “COVID-19 vaccination rate monitoring in Germany (COVIMO)”, was designed to be sampled on a monthly basis (75). From July 2021 until December 2021 and February until June 2022, continuous monthly interviews were carried out within the frameworks of GEDA 2021 and GEDA 2022 (82), respectively (see Figure 1).

The GEDA surveys and COVIMO were conducted on behalf of the Federal Ministry of Health of Germany. Data was collected by an external market and social research institute (USUMA GmbH). Study design, data collection, and sampling were largely the same across these different surveys, and based on a dual frame approach of mobile and landline numbers. The study population differs between the GEDA surveys and COVIMO, as GEDA targets people aged 15 or older living in private households whose main residence is in Germany (70), whereas COVIMO includes only people aged 18 or older (75). In addition, there is a slight difference in the general content focus of the studies (general health survey in GEDA versus vaccination monitoring in COVIMO). Details of the data pipeline, which cover semi-automated data preparation, data merging, and output creation as well as a description of sample weighting can be found elsewhere (83). We examined potential study-related differences between the GEDA surveys and COVIMO in the indicators of interest: Distributions of outcome variable scores were compared between studies in their one overlapping month using boxplots and violin plots (results not shown). No pronounced differences were detected.

#### 2.1.2 Participants

Across the entire survey period, 45,102 participants aged 18 or older were included in the analyses. The distributions by gender, age, and level of education in the different surveys are shown in Table 1, number of monthly cases are shown in Figure 2. The GEDA-EHIS 2019, GEDA 2020, 2021, and COVIMO studies surveyed approximately 1,000 participants per month. GEDA 2022 provided data for 3,000 participants per month. To reduce data gaps, the second half of each month was combined with the first half of the following month across the observation period.

**Table 1:**
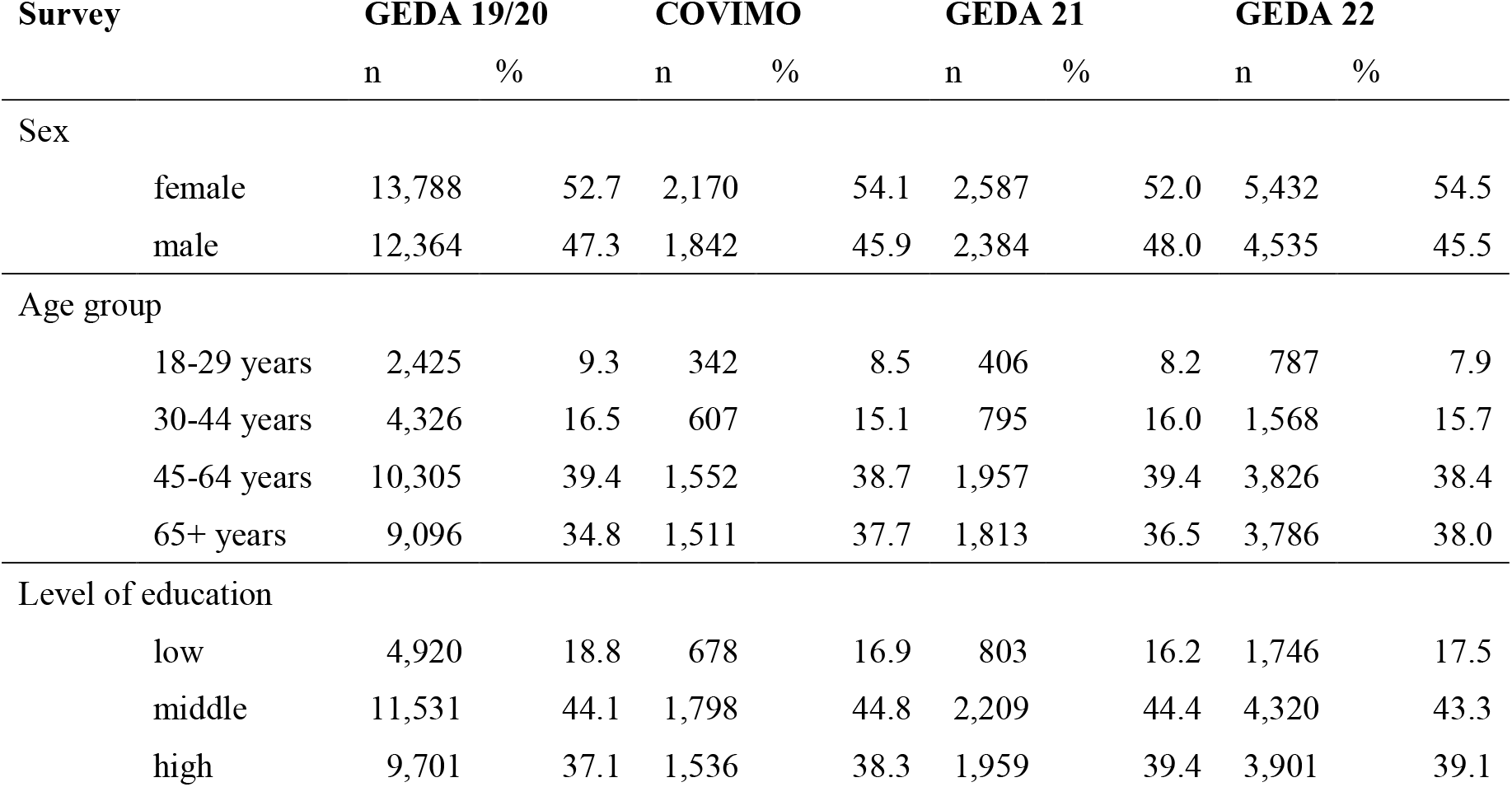
Sample composition

| Survey | GEDA 19/20 |  | COVIMO |  | GEDA 21 |  | GEDA 22 |  |
| --- | --- | --- | --- | --- | --- | --- | --- | --- |
|  | n | % | n | % | n | % | n | % |
| Sex |  |  |  |  |  |  |  |  |
| female | 13,788 | 52.7 | 2,170 | 54.1 | 2,587 | 52.0 | 5,432 | 54.5 |
| male | 12,364 | 47.3 | 1,842 | 45.9 | 2,384 | 48.0 | 4,535 | 45.5 |
| Age group |  |  |  |  |  |  |  |  |
| 18-29 years | 2,425 | 9.3 | 342 | 8.5 | 406 | 8.2 | 787 | 7.9 |
| 30-44 years | 4,326 | 16.5 | 607 | 15.1 | 795 | 16.0 | 1,568 | 15.7 |
| 45-64 years | 10,305 | 39.4 | 1,552 | 38.7 | 1,957 | 39.4 | 3,826 | 38.4 |
| 65+ years | 9,096 | 34.8 | 1,511 | 37.7 | 1,813 | 36.5 | 3,786 | 38.0 |
| Level of education |  |  |  |  |  |  |  |  |
| low | 4,920 | 18.8 | 678 | 16.9 | 803 | 16.2 | 1,746 | 17.5 |
| middle | 11,531 | 44.1 | 1,798 | 44.8 | 2,209 | 44.4 | 4,320 | 43.3 |
| high | 9,701 | 37.1 | 1,536 | 38.3 | 1,959 | 39.4 | 3,901 | 39.1 |

**Figure 2:**
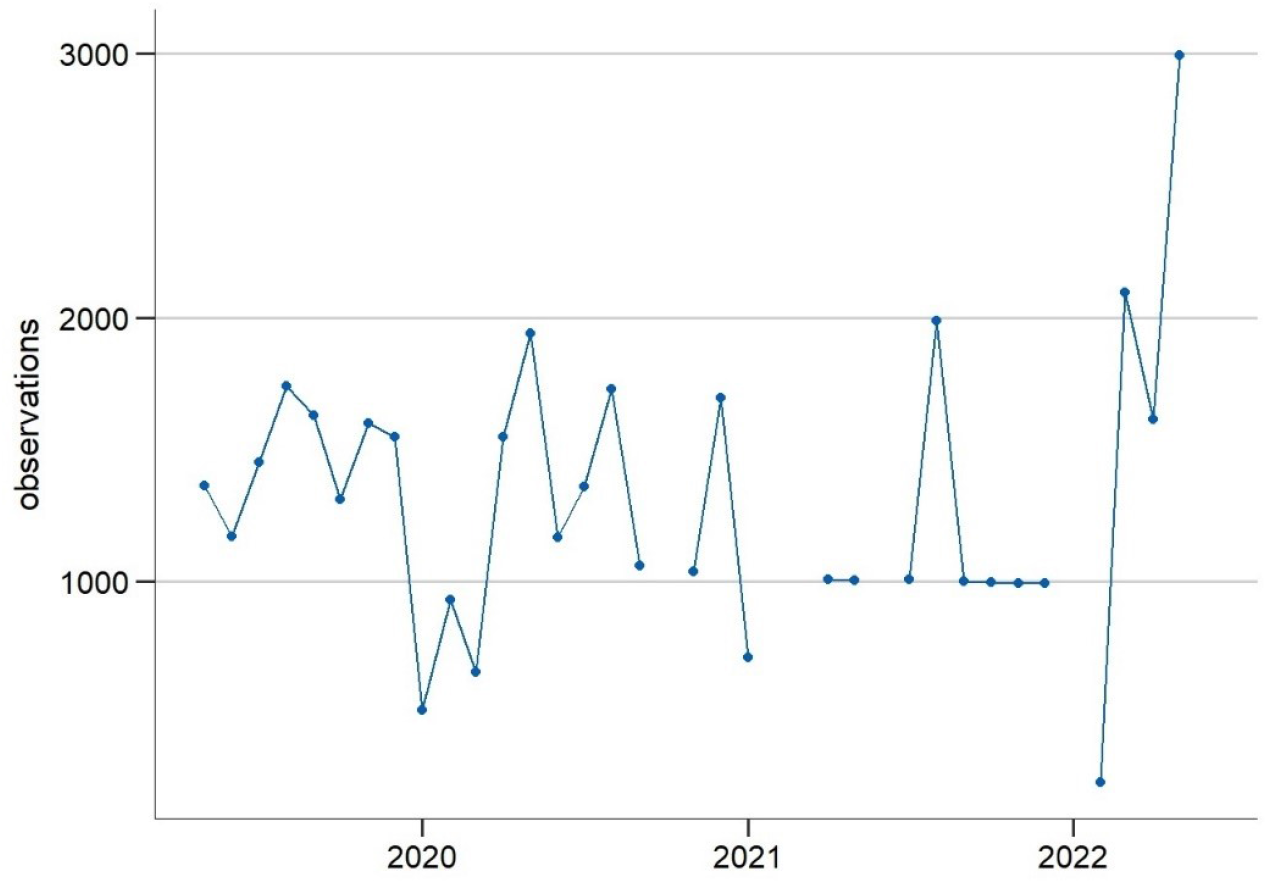
Observations per one-month period ranging from middle of the month to middle of the following month

### 2.2 Indicators of Mental Health Status

#### 2.2.1 Depressive symptoms

Depressive symptoms were observed prior to and during the pandemic using monthly data from beginning of April 2019 until mid-June 2022 (see Figure 1). There were four short data gaps (the largest between January and mid-March 2021). The indicator was measured with the established ultra-brief screening instrument “Patient Health Questionnaire-2” (PHQ-2) (78), which has been found to perform well as a screening tool for depressive disorders in the German general population (84). The PHQ-2 captures the frequencies of two core symptoms of depressive disorders, asking, “Over the last 2 weeks, how often have you been bothered by the following problems?”: 1) “little interest and pleasure in doing things” 2) “feeling down, depressed or hopeless” (possible responses: 0 = “not at all”, 1 = “several days”, 2 = “more than half the days”, 3 = “nearly every day”). The total score of the PHQ-2 ranges from 0 to 6 (‘no symptoms’ to ‘severe symptoms’). According to scoring recommendations (84), scores ≥ 3 represent a positive screen for possible depressive disorder and indicate a potential need for further diagnostic assessment. Two measures are reported in the current study: (1) the mean depressive symptom score, which tracks changes in the mean severity of symptoms in the population (65); (2) the proportion of the adult population screening positive for possible depressive disorder.

#### 2.2.2 Symptoms of anxiety

Symptoms of anxiety were observed monthly from mid-March 2021 to mid-June 2022 (see Figure 1) with two short data gaps (mid-May 2021 to mid-June 2021 and January 2022). The indicator was measured with the established ultra-brief screening instrument “Generalized Anxiety Disorder-2” (GAD-2), which has been found to perform well as a screening tool for anxiety disorders in the German general population (80). The GAD-2 captures the frequency of two core symptoms of anxiety disorders, asking, “Over the last 2 weeks, how often have you been bothered by the following problems?”: 1) “feeling nervous, anxious or on edge” 2) “ not being able to stop or control worrying” (possible responses: 0 = “not at all”, 1 = “several days”, 2 = “more than half the days”, 3 = “nearly every day”). The total score of the GAD-2 ranges from 0 to 6 (no symptoms to severe symptoms). Scores ≥ 3 represent a positive screen for possible anxiety disorder, including generalized anxiety disorder, panic disorder, social anxiety disorder, and posttraumatic stress disorder (80). Just as with depressive symptoms, two measures are reported: (1) the mean anxiety symptom score and (2) the proportion of the adult population screening positive for possible anxiety disorder.

#### 2.2.3 Self-rated Mental Health (SRMH)

SRMH was observed monthly from mid-March 2021 to mid-June 2022 (see Figure 1) with two short data gaps (mid-May 2021 to mid-June 2021 and January 2022). It was measured using the question: “How would you describe your overall mental health?” (possible responses: 5 = “excellent”, 4 = “very good”, 3 = “good”, 2 = “fair”, 1 = “poor”). The single item is an established way to measure SRMH in population based surveys (81). SRMH has been found to represent a dimension of mental health that is qualitatively distinct from psychopathology (85). Here and elsewhere (50) it is employed as a measure of positive mental health. Two measures are reported: (1) population mean SRMH score; (2) the proportion of the adult population rating their mental health as “very good” or “excellent”, following previous categorization to identify the presence of positive mental health (50).

#### 2.3 Sociodemographic variables used to measure mental health inequalities

Results are presented separately for women and men. For this purpose, respondents’ information on the sex noted in their birth certificate was used. Information on gender could not be used in the present analyses, since the data for the evaluations are adjusted to the marginal distributions of the official reference statistics (source: Microcensus (86)), which lacks information on gender identity.

Four age groups were formed to capture young adulthood, different stages of middle age, and the ages of an increased risk of severe COVID-19 infection: 18 to 29 years, 30 to 44 years, 45 to 64 years, and 65 years and older.

Educational levels according to the CASMIN classification (“Comparative Analyses of Social Mobility in Industrial Nations”) were used as an indicator of socioeconomic status (87). Three groups with low, medium, and high levels of education are distinguished on the basis of school and vocational qualifications.

### 2.4 Statistical Analysis

All analyses were conducted in R version 4.1.2 and Stata /SE 17.0.

#### 2.4.1 Estimation of moving three-month averages and smoothing curves

In order to assess mental health developments over time in the general population and by subgroup, we calculated time series of estimates along with smoothing curves to be represented graphically (for details, see (83)). Our aim was to achieve high temporal resolution whilst working with sample size restrictions and also to smooth random fluctuations. The estimation procedure described below also ensures that possible fluctuations in distributions of sex, age, and level of education in the sample over time are corrected for and that stratified results are standardized for the other main sociodemographic characteristics.

For each of the three mental health indicators, linear and logistic regressions were used to predict a time series of means and proportions for the adult population in Germany. To handle low cell counts and reduce volatility over time, we estimated centered moving averages rather than monthly averages (88) using weighted data from three-month windows. Some three-month windows only included data from two months due to data gaps. The three-month windows move in steps of one month. The models for each three-month window regress the mental health indicators on sex, age group, and level of education, and interactions between them. While the linear models include all possible interaction terms, only all possible two-way interactions of the covariates but not the three-way interactions were included in the logistic regression models to avoid problems resulting from empty cells. Nonetheless, there were some empty cells, resulting in estimation gaps in the time series of categorical PHQ-2 and GAD-2 estimates.

These regression models are the foundation for standardization for sex, age, and level of education between the three-month windows, which ensures that different distributions of these characteristics between them do not influence the results. For standardization, we calculated averaged predictions in a two-step process. First, we used the models to perform predictions on a standard population. To calculate arithmetic means of mental health indicator scores, we used the model estimates from the linear regressions and predicted the expected values of the indicator in question. To calculate proportions for categorical indicator outcomes, we predicted the expected probabilities. In a second step we averaged over all of the predictions. The standard population was calculated using data from the Microcensus 2018 (86), which approximates Germany’s population in 2018.

The calculation of estimates for time series stratified by sex, age, and level of education was similar to the procedure described above. However, in order to exclude different distributions of the respective other two characteristics in different time periods as explanatory factors for temporal developments, stratified results by age group, sex, or level of education were standardized by the remaining two characteristics in the prediction step. For example, the results stratified by age group were standardized for sex and education. This was achieved by making predictions for every subgroup as if all observations in the standard population belonged to this subgroup. The standardization between subgroups means that the subgroup-specific estimates are not representative for the population subgroup. The mathematical and methodological foundations for model-based predictions and standardization can be found elsewhere (89-91).

In order to improve results interpretation by making trends more visible, we additionally estimated smoothed curves using a general additive model (92) with a smoothing spline (93, 94) and curve by factor interaction (95). Values were predicted on the same standard population. The spline was fitted on weekly observations to maximize temporal resolution given sample size. To avoid over- or underfitting, the smoothing parameter was estimated using restricted maximum likelihood. However, we found that for our shorter time series, the curves based on weekly estimates were less smooth than the three-monthly predictions. Therefore, we only used this procedure for the longer time series.

Missing values in the dependent variables were excluded on a case-by-case basis. Observations without information on sex or age were not included in the survey and were treated as non-responses. Missings in education were imputed in accordance with the weighting procedure (66) by assigning the most frequent value, a medium level of education.

The initial interpretation of the curves was descriptive by visual inspection. Conservative criteria such as confidence interval comparisons were not used to evaluate developments over time at this stage because the first aim was to explore the overall trajectory. In addition to visual inspection, we carried out statistical comparisons between different time periods (see 2.4.2).

#### 2.4.2 Statistical time period comparisons between survey years

For the longer depressive symptoms time series, we conducted statistical comparisons between the three survey years for two periods of months: 1) mid-March to mid-September (calendar week (CW) 11-37) 2019, 2020, 2021, mid-March to mid-June (CW 11-24) 2022, and 2) mid-September to end of December 2019, 2020, 2021 (CW 38-52). For the shorter time series of SRMH and anxiety symptoms, mid-March to mid-September 2021 (CW 11-37) and mid-March to mid-June (CW 11-24) 2022 were compared. These time periods were chosen based on (1) the declaration of a pandemic on March 11 from WHO (1), (2) data gaps for the initial months of each year, and (3) the turning point in the development of depressive symptoms at the end of the summer of 2020, as shown in Figure 3 below. Small gaps in some of these time periods could not be avoided; for example, there was no data from mid-March to the beginning of April 2019. We tested for two kinds of trends. First, we tested for differences in mean depressive or anxiety symptom score and proportions at or above PHQ-2/ GAD-2 cutoff between corresponding time periods across years for the overall population as well as within the different subgroups. For SRMH we tested for differences in mean and proportions with very good or excellent SRMH. Second, we tested for possible changes in differences in means or proportions between subgroups over the specified time periods. Individual groups were tested against each other only in the case of an overall significant difference.

**Figure 3:**
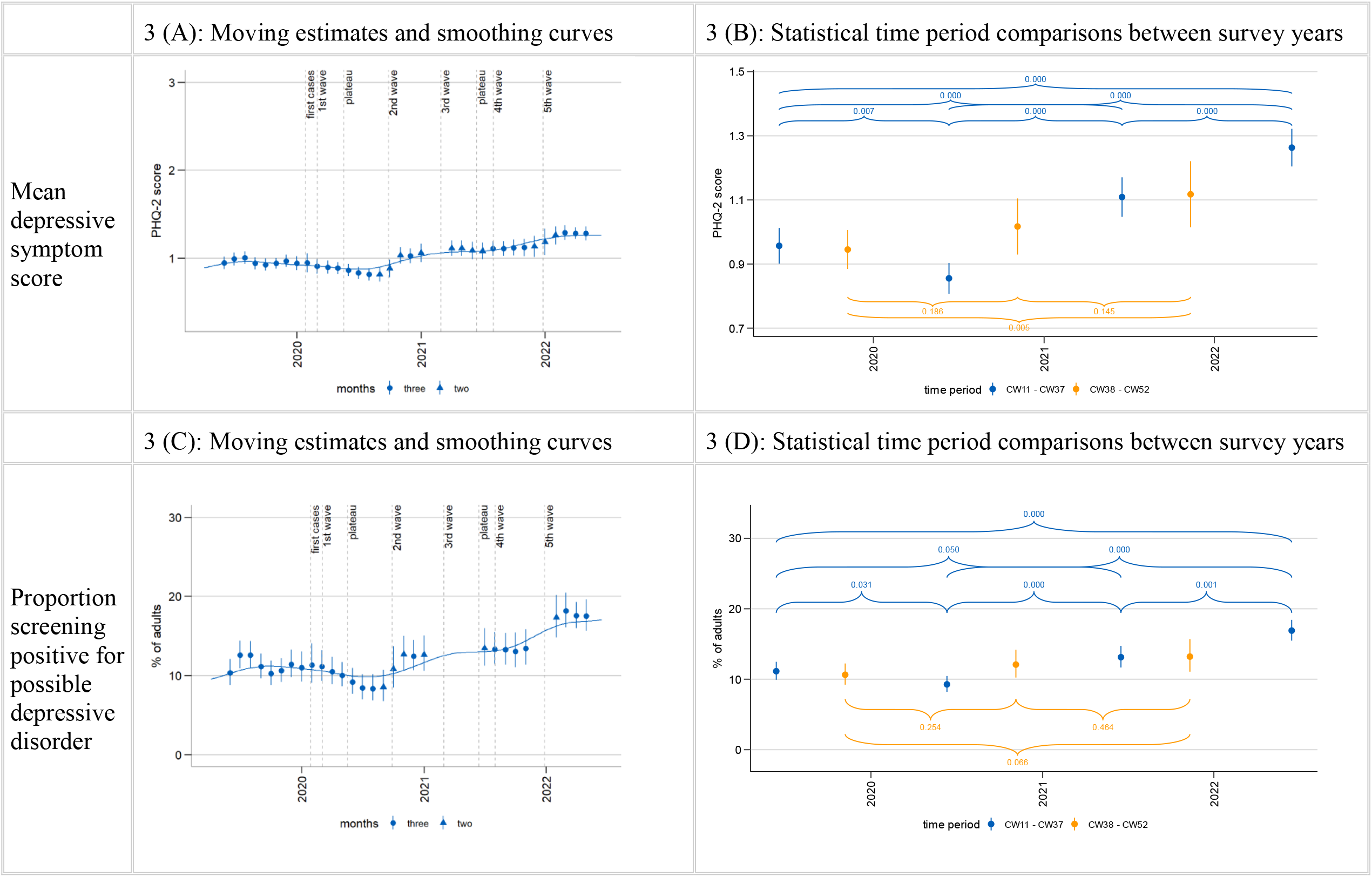
Time trends in depressive symptoms

To conduct these comparisons, we again used linear and logistic regression models to produce averaged predictions as described above. However, here we calculated estimates for the defined time periods by including a set of dummy variables indicating these periods in the survey year. Furthermore, all possible interactions of these dummies with age group, sex, and level of education were included. The specification of the linear and the logistic regression models again differed with regard to level of interaction. In the logistic regression only, three-way interactions were included, whereas the linear model also included four-way interactions. After model estimation, the standard population was used for prediction of the means of the specified time periods. Contrasts between the time periods and the differences between the subgroups between the time periods were estimated. We used Stata’s ‘margins, contrast’ command (96) for estimation and statistical testing using Wald tests, applying a significance level of (p<0.05).

## 3 Results

### 3.1 Time-trends of mental health indicators in the total population

#### 3.1.1 Depressive symptoms

Symptoms of depression were observed from April 2019 to June 2022. As shown in Figure 3 (A,C), the estimated population means of the PHQ-2 ranged from 0.81 (95 % CI: 0.73 – 0.90) to 1.29 (95 % CI: 1.21 – 1.37) points in the observation period, well below the screening cutoff (score ≥ 3). The percentage of those screening positive for possible depressive disorder ranged from 8.4 % (95 % CI: 6.9 % – 10.1 %) to 18.2 % (95 % CI: 16.1 % – 20.4 %). Visual inspection of the moving three-month averages suggests that the mean depressive symptom scores in the general population as well as the proportion of the population with a positive screen decreased during the first wave of the pandemic and into the first summer plateau in 2020. Statistical comparisons between specific time periods confirm this finding (see Figure 3 (B,D)): both mean scores and proportions at or above cutoff were significantly lower in the months after the outbreak of the pandemic from mid-March to mid-September 2020 than in the same period before the pandemic in 2019.

After the first summer plateau, the time series are characterized by two increases: Both PHQ-2 measures (means and positive screens) first increased between the beginning of the second wave in autumn 2020 and the beginning of the third wave in spring 2021. They reached relatively steady levels above 2019 between spring 2021 and autumn 2021 (see significant differences for both measures between mid-March to mid-September 2019 and 2021 and for means between-September to end-December 2019 and 2021 in Tables 2,3,4 and Figure 3 (B,D)). Both then showed further increases from late 2021 to February/March 2022, particularly between the last 2021 and the first 2022 estimates, and remained elevated until the end of the observation period. Spring/summer 2022 depressive symptom levels were significantly higher than in the same months in all three previous years (see significant differences between mid-March to mid-September 2019, 2020, and 2021 and mid-March to mid-June 2022).

**Table 2:** Depressive symptoms in different time periods: mean symptom scores (PHQ-2)

|  | cw 11-37 2019 |  |  | cw 38-52 2019 |  |  | cw 11-37 2020 |  |  | cw 38-52 2020 |  |  | cw 11-37 2021 |  |  | cw 38-52 2021 |  |  | cw 11-24 2022 |  |  |
| --- | --- | --- | --- | --- | --- | --- | --- | --- | --- | --- | --- | --- | --- | --- | --- | --- | --- | --- | --- | --- | --- |
|  | mean | 95%-CI |  | mean | 95%-CI |  | mean | 95%-CI |  | mean | 95%-CI |  | mean | 95%-CI |  | mean | 95%-CI |  | mean | 95%-CI |  |
| **Total** | **0.96** | **0.90** | **1.01** | **0.95** | **0.88** | **1.01** | **0.86** | **0.81** | **0.90** | **1.02** | **0.93** | **1.10** | **1.11** | **1.05** | **1.17** | **1.12** | **1.01** | **1.22** | **1.26** | **1.20** | **1.32** |
| **Sex** |  |  |  |  |  |  |  |  |  |  |  |  |  |  |  |  |  |  |  |  |  |
| male | **0.93** | 0.85 | 1.01 | **0.92** | 0.83 | 1.01 | **0.78** | 0.71 | 0.85 | **0.95** | 0.83 | 1.08 | **1.00** | 0.92 | 1.09 | **1.01** | 0.87 | 1.16 | **1.19** | 1.10 | 1.28 |
| female | **1.00** | 0.92 | 1.08 | **0.98** | 0.89 | 1.07 | **0.94** | 0.87 | 1.01 | **1.08** | 0.96 | 1.21 | **1.22** | 1.13 | 1.32 | **1.23** | 1.07 | 1.38 | **1.33** | 1.26 | 1.41 |
| **Age** |  |  |  |  |  |  |  |  |  |  |  |  |  |  |  |  |  |  |  |  |  |
| 18-29 | **1.12** | 0.91 | 1.33 | **1.01** | 0.80 | 1.22 | **1.10** | 0.95 | 1.26 | **0.95** | 0.74 | 1.16 | **1.30** | 1.06 | 1.54 | **1.57** | 1.04 | 2.10 | **1.35** | 1.18 | 1.52 |
| 30-44 | **0.89** | 0.75 | 1.03 | **1.09** | 0.89 | 1.30 | **0.77** | 0.64 | 0.89 | **1.07** | 0.77 | 1.38 | **1.21** | 1.02 | 1.40 | **1.00** | 0.80 | 1.20 | **1.09** | 0.95 | 1.23 |
| 45-64 | **1.09** | 0.98 | 1.19 | **1.03** | 0.93 | 1.14 | **0.91** | 0.82 | 1.00 | **1.11** | 0.96 | 1.27 | **1.07** | 0.96 | 1.17 | **1.17** | 0.99 | 1.35 | **1.37** | 1.26 | 1.47 |
| 65+ | **0.80** | 0.71 | 0.89 | **0.69** | 0.61 | 0.77 | **0.73** | 0.65 | 0.81 | **0.86** | 0.73 | 0.99 | **0.94** | 0.86 | 1.02 | **0.86** | 0.72 | 1.00 | **1.15** | 1.07 | 1.24 |
| **Level of education** |  |  |  |  |  |  |  |  |  |  |  |  |  |  |  |  |  |  |  |  |  |
| low | **1.20** | 1.05 | 1.35 | **1.12** | 0.96 | 1.27 | **1.02** | 0.91 | 1.14 | **1.15** | 0.92 | 1.38 | **1.33** | 1.16 | 1.50 | **1.31** | 1.01 | 1.60 | **1.45** | 1.31 | 1.58 |
| middle | **0.91** | 0.84 | 0.98 | **0.94** | 0.85 | 1.03 | **0.81** | 0.75 | 0.87 | **1.01** | 0.90 | 1.12 | **1.02** | 0.94 | 1.09 | **1.06** | 0.95 | 1.18 | **1.20** | 1.12 | 1.27 |
| high | **0.68** | 0.62 | 0.74 | **0.71** | 0.63 | 0.79 | **0.66** | 0.58 | 0.74 | **0.77** | 0.68 | 0.85 | **0.87** | 0.81 | 0.94 | **0.88** | 0.77 | 0.99 | **0.98** | 0.92 | 1.04 |

**Table 3:**
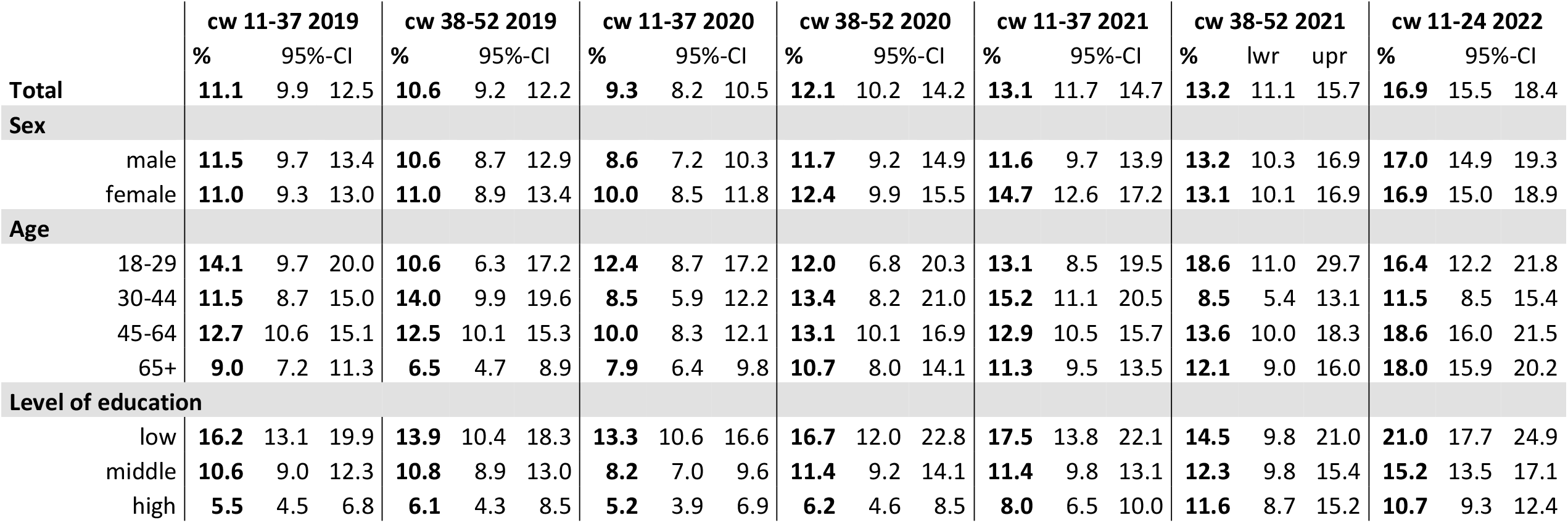
Depressive symptoms in different time periods: percentages of positive screens (PHQ-2 score > 2)

**Table 4:**
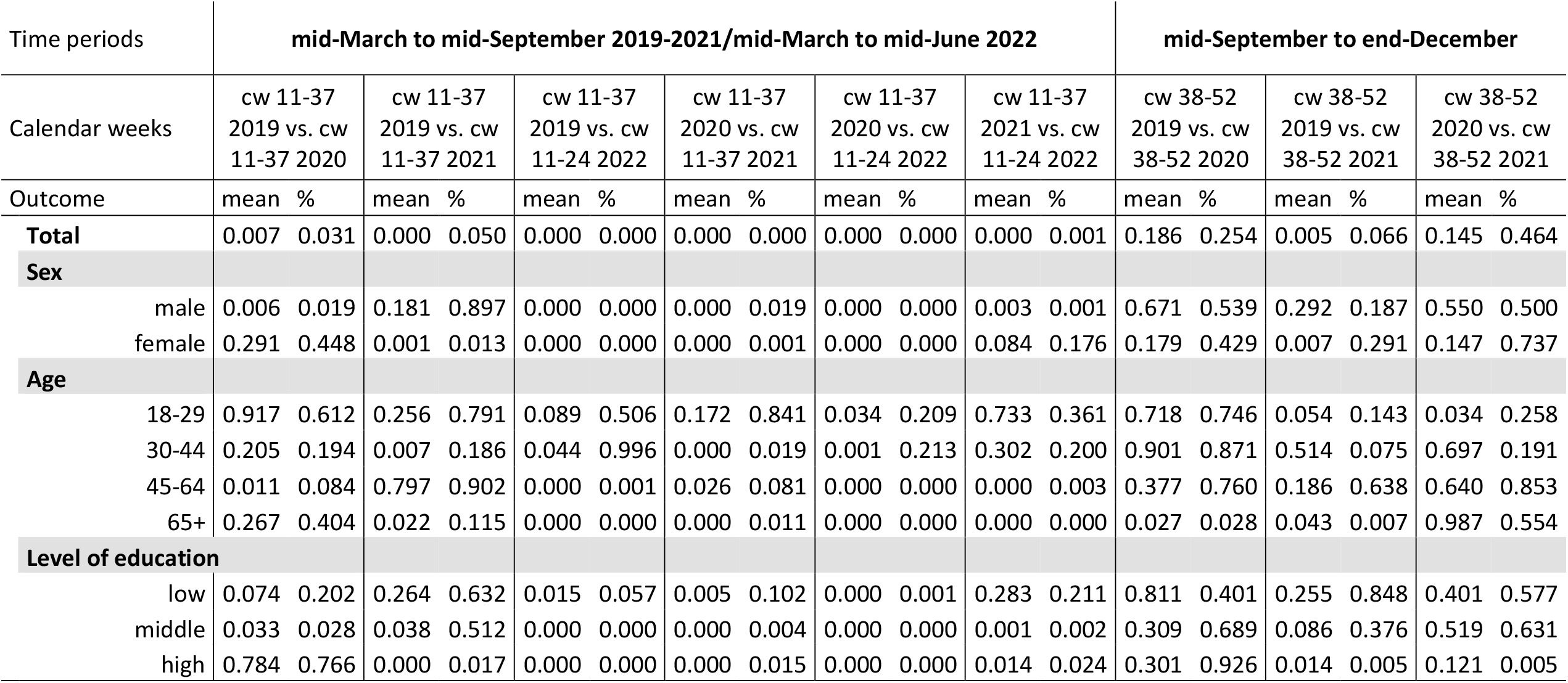
Depressive symptoms in different time periods: p-values for statistical comparisons

This overall trajectory manifests in the following development of time period estimates: The mean depressive symptom score in the population first decreased from steady levels of 0.96 and 0.95 in mid-March to mid-September and mid-September to end of December 2019 to 0.86 in mid-March to mid-September 2020. It then increased to 1.11 and 1.12 in spring/summer and autumn/winter 2021, respectively, (see Table 2) and to 1.26 in mid-March to mid-June 2022. The proportion of the population with a positive screen (see Table 3) first decreased from 11.1 % in the period from April to mid-September 2019 to 9.3 % in mid-March to mid-September 2020. By mid-March to mid-September 2021, this proportion had risen to 13.1 %. Between mid-March and mid-June 2022, it averaged 16.9 %.

#### 3.1.2 Symptoms of anxiety and self-rated mental health (SRMH)

Symptoms of anxiety were observed from March 2021 to June 2022. As shown in Figure 4(C), the estimated population means of the GAD-2 ranged from approximately 0.66 (95 % CI: 0.60 – 0.72) to 0.96 (95 % CI: 0.89 – 1.03) points during the observation period, well below the screening cutoff. The percentage of those screened positive for possible anxiety disorder ranged from approximately 7.7 % (95 % CI: 5.9 % – 9.9 %) to 11.7% (95 % CI: 10.0 % – 13.7 %) (see Figure 4(D)). The moving averages suggest a possible increase in mean anxiety score in the population from spring 2021 into autumn 2021, flattening out by the end of 2021 (see Figure 4(C)). This development is hardly reflected in the proportion of those exceeding the cut-off value for possible anxiety disorder; however, empty cells made it impossible to calculate the first and last estimates of 2021 for the categorical outcome. With the first estimate in 2022, the moving averages then showed a marked increase in mean symptom scores and proportions of positive screens and consistently elevated levels until the end of the observation period. Statistical time period comparisons confirm an increase between spring/summer 2021 and 2022 (see Table 5). Symptoms of anxiety increased from a population mean score of 0.75 and a proportion of positive screens of 7.2 % in mid-March to mid-September 2021 to 0.96 and 11.1 %, respectively, in mid-March to mid-June 2022.

**Figure 4:**
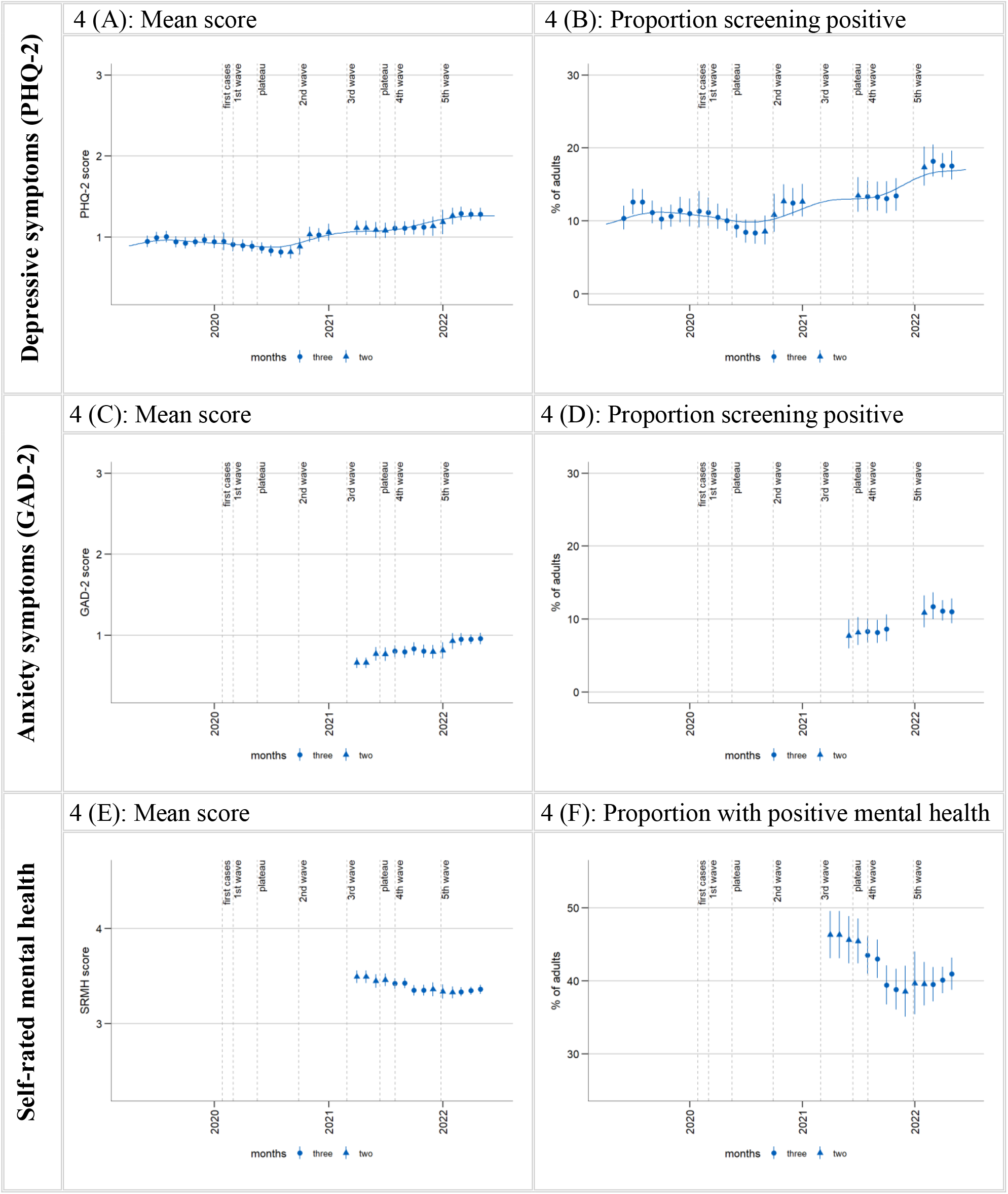
Time trends in depressive symptoms, anxiety symptoms, and SRMH

**Table 5:**
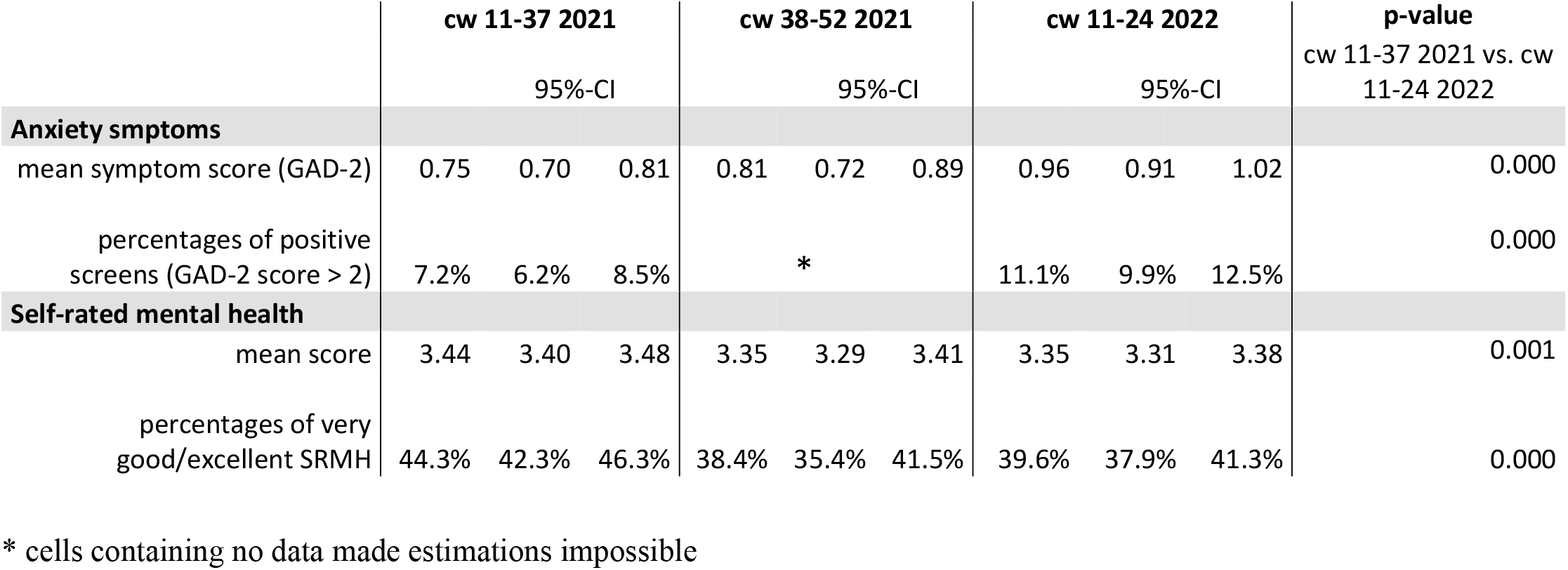
Anxiety symptoms and self-rated mental health in different time periods: mean values, percentages and p-values for statistical comparisons

SRMH was observed from March 2021 to June 2022. As shown in Figure 4(E), estimated population means of SRMH ranged from 3.33 (95 % CI: 3.27 – 3.39) to 3.50 (95 % CI: 3.42 – 3.56) points in the observation period, i.e. from good (3 points) to very good (4 points) SRMH. As shown in Figure 4(F), the percentage of those with very good or excellent SRMH ranged from 38.5 % (95 % CI: 35.1% – 42.0%) to 46.3 % (95 % CI: 43.1 % – 49.5%). The plotted time series show a nearly steady decline in both mean SRMH and the percentage of those with very good or excellent SRMH between spring 2021 and autumn 2021. Mean SRMH remained at a fairly constant level for the rest of the observation period. However, the percentage of those with very good or excellent SRMH continued to decline until the end of 2021 and then increased in spring/early summer 2022. The overall declines in both mean SRMH and the percentage of the population with very good or excellent SRMH in the course of the observation period were confirmed by statistical time period comparisons (see significant differences between spring/summer 2021 and 2022 in Table 5). SRMH declined from a population mean score of 3.44 and a proportion of those with very good or excellent SRMH of 44.3 % in mid-March to mid-September 2021 down to 3.35 and 39.6 %, respectively, in mid-March to mid-June 2022

In summary, looking at all three indicators for just this shorter time period, depressive and anxiety symptoms both increased significantly between spring/summer 2021 and spring/summer 2022, while SRMH declined. Symptoms of depression and anxiety showed marked increases between the final estimates of 2021 and the first estimates of 2022 and remained elevated, while SRMH showed no marked changes at this time and improved at the end of the observation period.

### 3.2 Differences in depressive symptoms according to sociodemographic characteristics

The subgroup estimates reported below are standardized values. They should not be taken as population estimates for these groups.

#### 3.2.1 Differences according to sex

Throughout the observation period, mean depressive symptom scores standardized for education and age were higher in women than in men (see Figure 5(A)). Likewise, standardized percentages of positive screens for possible depressive disorder were, for the most part, higher in women than in men, although the overlap was greater than for mean scores (see Figure 5(C)). The plotted time series indicate lower depressive symptom levels in men between the beginning of the first wave and the end of the summer plateau in 2020 and in women in the summer plateau. Statistical time period comparisons of mid-March to mid-September 2019 vs. 2020 confirm this trend only for men both regarding mean symptom score and proportion of those screening positive (see Figures 5(B,D), Tables 2,3,4). Both measures then showed an increase between the onset of the 2^nd^ wave and the onset of the 3^rd^ wave in both sexes. Among men, they remained at a steady higher level which was not significantly elevated compared to pre-pandemic levels until the end of 2021 (see non-significant results 2019 vs. 2021). Among women, mean scores and, to a lesser extent, proportions of positive screens showed another increase from mid-2021 through the end of the year. Mean score estimates for women were significantly higher in both tested time periods in 2021 compared to pre-pandemic periods (see Figure 5(B), Tables 2,3,4). Proportions of women screening positive showed a significant increase in mid-March to mid-September 2021 compared to 2019. For 2022, Figures 5(A,C) show an initial continuation of increases in mean symptom scores and proportions of positive screens in women followed by a slight decline in both in the final two estimates in the time series.

**Figure 5:**
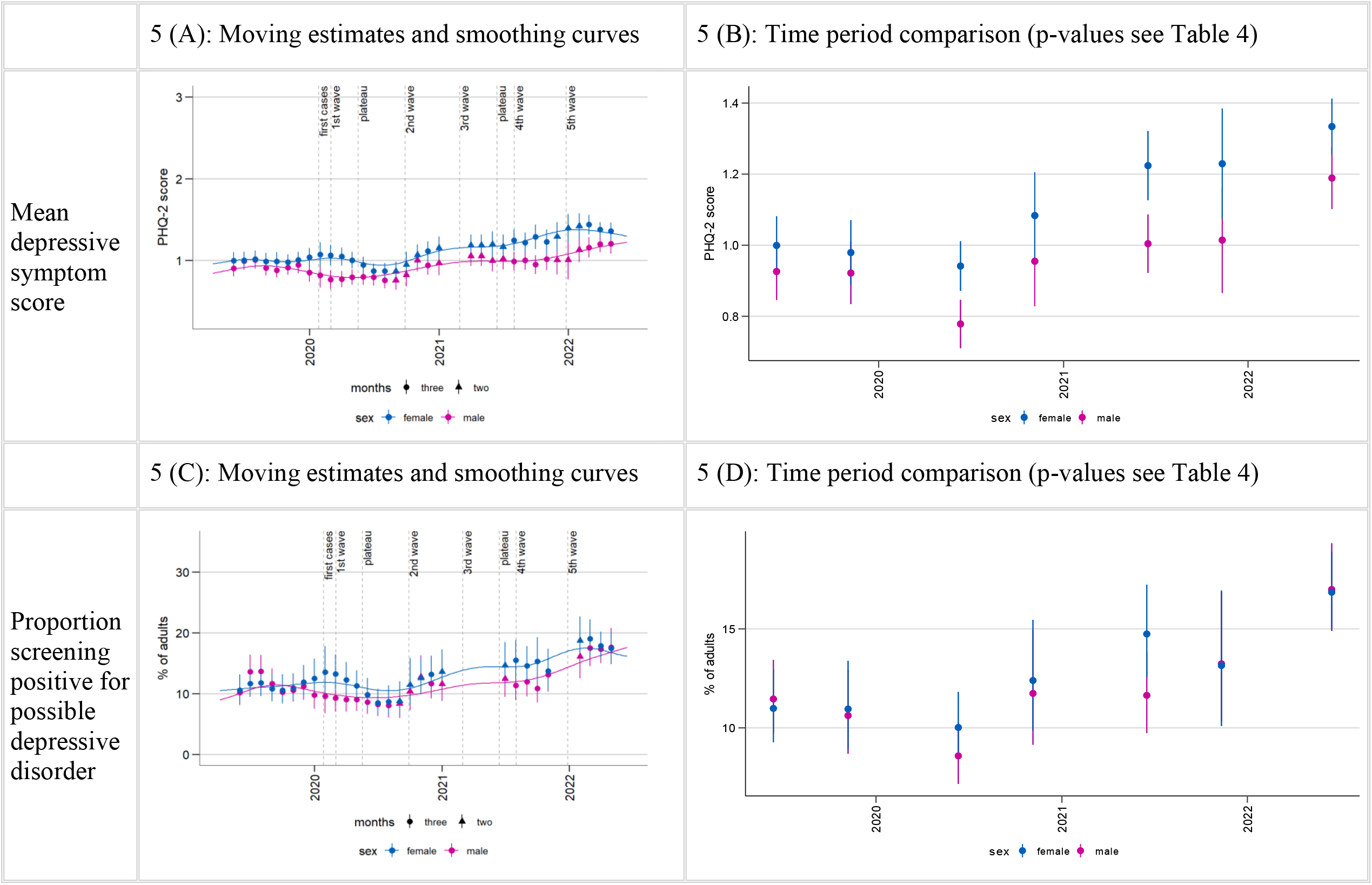
Time trends in depressive symptoms by sex (standardized by age, level of education)

Men, who showed fairly constant depressive symptom levels for most of 2021, experienced a substantial increase in both PHQ-2 outcomes in early 2022, followed by a continued upward trend until the end of the time series. Correspondingly, 2022 increases do not quite reach statistical significance in women but do in men (see comparisons mid-March to mid-September/mid-June 2021 vs. 2022 in Tables 2,3,4). With these increases, men significantly surpassed pre-pandemic levels of mean symptom scores and positive screens for the first time, and women remained significantly above 2019 levels (comparisons mid-March to mid-September/mid-June 2019 vs. 2022 in Table 4). Among women, mean scores and the percentage of positive screens standardized by age and level of education increased from 1.00 and 11.0 % in spring/summer 2019, respectively, to 1.33 and 16.9 % in spring/summer 2022 in the course of the observation period; for men from 0.93 and 11.5 % to 1.19 and 17.0 %. While these trends suggest that the differences in depressive symptoms between women and men might have changed in the course of the time series, a joint test showed no significant dependency of the difference between the sexes on time period (results not shown). Therefore, pairwise comparisons were not evaluated.

#### 3.2.2 Differences according to age

Across the whole observation period, those aged 65+ years tended to exhibit lower mean symptom scores and those aged 18-29 years tended to exhibit higher mean symptom scores than the other age groups (all age groups standardized by sex and education; see Figure 6(A)). This pattern is less pronounced in the proportions of positive screens time series (see Figure 6(C)). Among all age groups except in those aged 65+, visual inspection reveals a decline in mean symptom scores as well as in the proportions of those screening positive in the initial phases of the pandemic. However, these declines resulted in significant differences between the mean values from mid-March to mid-September 2020 compared with 2019 only for those aged 45-64 years. While depressive symptoms increased briefly among 18-to 29-year-olds with the outbreak of the pandemic and did not decline until the summer months, they declined from the beginning of the outbreak in the two middle age groups. At different times between the second wave and the end of the observation period, every age group then showed increases in depressive symptom levels beyond pre-pandemic levels.

**Figure 6:**
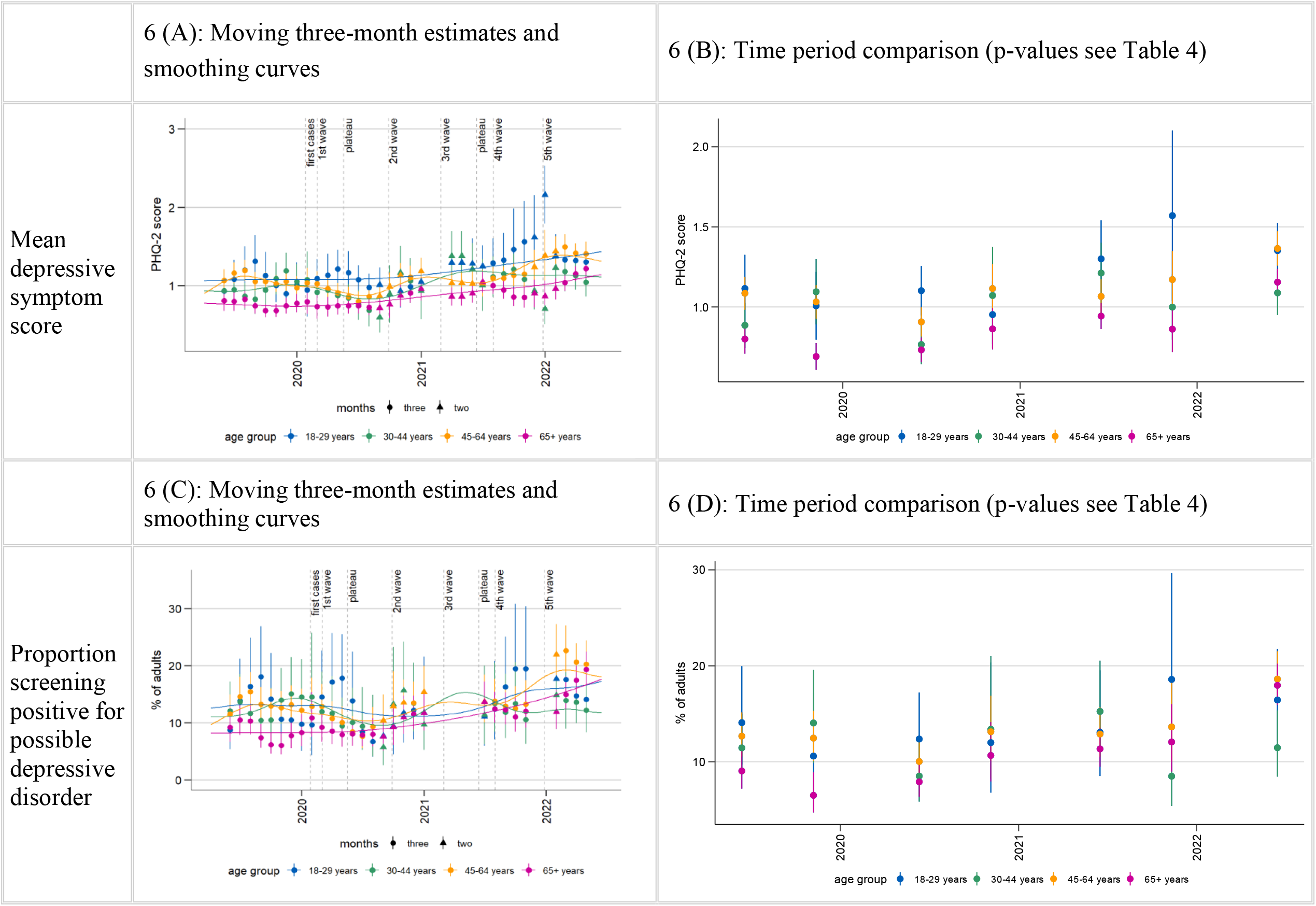
Time trends in depressive symptoms by age groups (standardized by sex, level of education)

Despite substantial fluctuations in the moving average estimates, smoothing splines among 18-to 29-years-olds suggest an overall increase from late 2020/ early 2021 until the end of the observation period in mean values and proportions of positive screens. Both estimates rose in the first half of 2021 and very markedly compared to other groups from fall 2021 to the end of the year, with means reaching higher levels than in any other group at any other time. The standardized proportion of positive screens rose from 10.6 % between mid-September to end of December 2019 to 18.6 % for the same period in 2021 (see Figure 6(D)). However, in this age group, these increases did not reach significance compared to pre-pandemic levels (see Tables 2,3,4). From 2022 onward, the monthly estimates declined back to 2021 levels for the same months.

30-to 44-year-olds showed a temporary increase in both PHQ-2 measures in spring/summer 2021. Standardized mean symptom scores increased significantly from 2019 in this time and markedly compared to other groups (from 0.89 in 2019 to 1.21 in 2021). The increase from 11.5 % to 15.2 % in the standardized proportion of positive screens does not reach statistical significance. Both measures then declined to 2019 levels in the later months of 2021. After a sudden increase in the early months of 2022 followed by a decline until the end of the observation period, mean symptom scores again significantly surpassed 2019 levels for the same months in mid-March to mid-June 2022, but not 2021 levels. However, proportions of positive screens did not show increases in this final observed time period.

45-to 64-year-olds exhibited an increase in depressive symptom levels above pre-pandemic levels later than the other age groups. The time series graphs show increases from about mid-2021, mostly in mean symptom scores. Both PHQ-2 measures significantly surpassed 2019 levels for the first time in 2022, also surpassing 2021 levels (comparison mid-March to mid-September/mid-June 2019 and 2021 versus 2022), with the standardized proportion of positive screens increasing from 12.7 % in spring/summer 2019 to 18.6 % in the same period in 2022 (see Figure 6(D)).

As indicated by the smoothing curve, 65+-year-olds exhibited an overall continuous increase in symptoms of depression following relatively stable mean values and positive screens until the summer plateau in 2020. Statistical time period comparisons also point to a particularly steady trend of increase in this age group. We found significant increases in both mean symptom scores and proportions of positive screens as early as mid-September to end-December in 2020 compared to 2019, in mean symptom scores in mid-March to mid-September 2021 compared to 2019, and in both outcomes in mid-September to end-December in 2021 compared to 2019. In 2022, this group experienced another marked increase that was ongoing until the end of the observation period, resulting in mean scores and proportions significantly above comparison periods in all previous years in mid-March through mid-June (see Tables 2,3,4), with the standardized proportion of positive screens reaching 18.0 % (compared to 2019: 9.0 %; 2020: 7.9 %; 2021: 11.3 %). A joint test showed no significant dependency of the difference between age groups on time period (results not shown). Therefore, pairwise comparisons to test changes in differences over time were not evaluated.

#### 3.2.3 Differences according to level of education

A social gradient is apparent throughout the observation period, with higher mean scores and proportions of positive screens for possible depression in those with the lowest levels of education, followed by the middle and high-level groups (all standardized by sex and age; see Figure 7(A,C)). In the early pandemic months of mid-March to mid-September 2020 time series plots for mean symptom scores as well as the proportion of positive screens indicate reductions in all groups as compared to the same time period in 2019. However, this decline was only transient and of small magnitude in the high level of education group. While the statistical comparison between spring/summer 2019 and 2020 reaches statistical significance in the middle level of education group, with proportions of positive screens decreasing from 10.6 % in 2019 to 8.2 % 2020, the decrease in the lowest level of education group from 16.2 % to 13.3 % does not reach statistical significance (see Table 3,4, Figure 7(D)).

**Figure 7:**
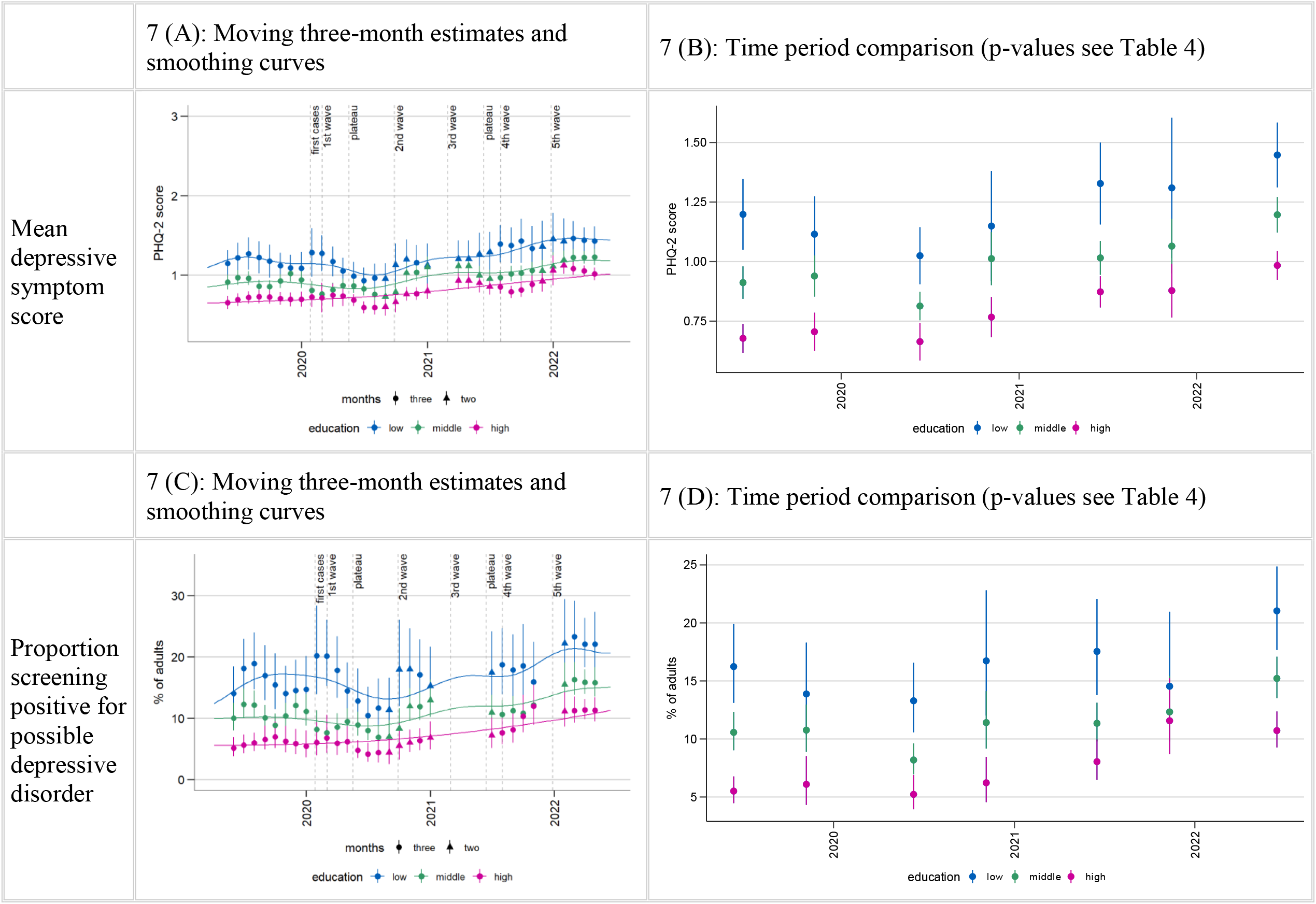
Time trends in depressive symptoms by level of education (standardized by age, sex)

The subsequent estimates show an overall trend of increases in mean symptom scores and proportions of positive screens between the 2^nd^ and 3^rd^ waves and another increase from mid-2021 until early 2022 in all groups. Smoothing splines and statistical comparisons suggest that this trend of an increase in both PHQ-2 outcomes was particularly continuous over time in individuals with a high level of education. This results in significantly higher mean scores and proportions of positive screens in both time period comparison windows in 2021 versus 2019 as well as in mid-March to mid-June 2022 compared to mid-March to mid-September 2021. Across the whole observation period, there was an increase from 5.5 % (mid-March to mid-September 2019) to 10.7 % standardized positive screens in about the same period of 2022. Smoothing splines for the other education groups suggest more fluctuation over time. Neither group showed significant increases above 2019 levels in 2021. However, the middle group did experience significant increases in both mean scores and proportions of positive screens between 2019 and 2022 as well as between 2021 and 2022. Increases in the low level of education group also reached significance compared to 2019 (but not 2021) in 2022, but only in mean symptom scores and not quite in the percentage of positive screens. The standardized proportions of those with a positive screen in the middle education group rose from 10.6 % in 2019 (mid-March to mid-September) to 15.2 % in 2022 (mid-March to mid-June) and from 16.2 % in 2019 to 21.0 % in 2022 in the low level of education group. A joint test showed no significant dependency of the difference between education groups on time period (results not shown). Therefore, pairwise comparisons to test changes in differences over time were not evaluated.

## 4. Discussion

### 4.1 Summary

The present study investigated how depressive symptoms developed month-by-month between April 2019 and June 2022 in the adult population in Germany and whether trajectories differ by sex, age, and level of education. Moreover, it explored how symptoms of anxiety and SRMH developed in the shorter time window of March 2021 to June 2022. We found:

1. Mean population depressive symptom scores as well as proportions of the population screening positive for possible depressive disorder showed a decline in the first wave of the pandemic and into the first summer plateau compared to the same months the year prior. Percentages of positive screens declined from about 11 % in spring/summer 2019 to 9 % in 2020. During the second wave starting in October 2020, this proportion as well as mean scores increased and remained consistently elevated throughout most of 2021, even during the summer months. Late 2021 until early spring 2022 saw another increase in both measures and sustained higher levels until the end of the observation period. By spring/summer 2021, the prevalence of those screening positive increased to 13 %, between March and June 2022, it averaged approximately 17%.
2. The observed overall trends in the development of depressive symptoms are, for the most part, evident across the examined subgroups. However, declines and increases are more pronounced in some groups than in others and vary in time course. The reduction in depressive symptoms in 2020 is particularly pronounced in men, among the two middle age groups, and the middle level of education group. Increases in depressive symptoms from autumn 2020 onward were found in all groups. However, numerically striking or statistically significant increases compared to pre-pandemic periods in 2019 were reached at different times. Developments among women, the youngest and eldest adults, and the high level of education group stand out. No changes in differences between groups were found for any sociodemographic characteristic across the observation period.
3. In keeping with these developments in depressive symptoms, SRMH decreased and anxiety symptoms increased between spring 2021 and early summer 2022.

### 4.2 Reduction in symptoms of depression in the first phases of the pandemic

Contrary to warnings of a potential mental health crisis at the start of the pandemic (2-5), our depressive symptom time series using the PHQ-2 show an initial reduction in both mean depressive symptom scores and proportions of individuals with a positive screen among adults in Germany during a first wave of infections. This first wave was mild compared to some other countries (97), and in the first pandemic summer, restrictions were eased and case numbers very low (20, 25). Analyses using the longer PHQ-8 in the same GEDA-EHIS data also showed a temporary reduction in symptoms of depression in the population between April 2020 and August 2020 in a month-by-month time series of the proportion of positive screens (98).

Findings on mental health in the early pandemic from other data sources and other countries are very mixed. This might be due to heterogeneity in observation and comparison periods and national contextual differences (e.g. 30, 35). In contrast to our results, international reviews and meta-analyses point out that many studies did find increases in psychological distress and symptoms of mental illness in the earliest phases of the pandemic (29, 30, 32, 99, 100), including symptoms of depression (32, 99, 101). While many studies with longer observation periods reported a decline back to or almost back to pre-pandemic levels in the summer months of 2020 (30, 32, 100), symptoms of depression were sometimes found to remain elevated for longer than symptoms of anxiety (32, 101). Also contradicting our results, a large population-based cohort study in Germany found an increase in individual-level PHQ-9 mean scores during the first wave of the pandemic (53) and in population-level means and proportions of positive PHQ-9 screens from 7.1 % to 9.5 % between April and November 2020 (38). Likewise, a longitudinal study based on representative household panel data found an increase in proportions of positive PHQ-2 screens from April to June 2020 (13.8 %) compared to the year 2019 (9.6 %) (37). These differences in findings may be due to differences in survey design such as the panel structure in the other studies in contrast to the monthly random samples in the present study and switches in survey mode from face-to-face to telephone interviews during the pandemic in one of the surveys (37). Differences in overall survey focus and framing (e.g. general health survey versus surveys with a special focus on the pandemic) as well as the institutions conducting the survey also cannot be ruled out as contributing factors. A representative regional study also using single-stage random sampling found no changes in psychopathological symptoms during the first wave compared to a pre-pandemic baseline (102), and another nationwide study found no changes within the weeks of the first lockdown compared to the weeks before (103). Further in keeping with a picture of resilient populations in the first wave, continuous reductions in symptoms of depression within the first months of the pandemic were reported in a large-scale study in the UK (104), and an Irish population-based study found a significantly lower proportion of positive screens for depression in March to April 2020 than in February 2019 (105).

Our analyses do not permit conclusions about causal associations between pandemic developments and mental health developments, much less on possible reasons for any putative associations between the two. However, the context within which mental health developments take place and their temporal coincidence with societal developments warrant discussion. Possible benefits of a general and novel deceleration of life during lockdown in the relatively mild first wave and relief from a relatively quick return to near-normalcy in the first pandemic summer could be taken into consideration as potential factors playing into the dynamics we find. Benefits of deceleration as a potential explanation is supported by the fact that analyses based on the same data examining all depressive symptoms included in the PHQ-8 found a particular reduction in fatigue, loss of energy, and concentration difficulties, which are all closely linked to chronic stress (79, 98).

Stratification by subgroups shows that while there is evidence of symptom reduction in the first pandemic summer in all groups except adults aged 65 years and older, lower symptom levels in the first wave and statistically significant reductions in spring/summer 2020 compared with spring/summer 2019 were found in groups that may have experienced a particular deceleration of life: the middle-aged, who are typically particularly busy with the demands of paid and unpaid work, and men, who, for example, took on less additional childcare than women when childcare facilities closed, particularly in high-income countries (45). The middle level of education group and, somewhat less markedly, the low level of education group also exhibit this pattern. Several workplace-related factors may have played a mediating role in a possible association between educational attainment and mental health, e.g. significantly reduced working hours with or without financial compensation versus increased working hours or job loss and working from home (38).

### 4.3 Declines in mental health from the second wave onward

#### 4.3.1 Declines in mental health from the second wave onward in the general population

While most studies on mental health in the COVID-19 pandemic in Germany examine its first months only (35), our results shed light on the development of symptoms of depression in the adult population until June 2022 and reveal two increases. Consistent with our finding of increased mean depressive symptom scores as well as positive screens between the last months of 2020 and spring 2021, i.e. during the second wave of infections, a German study reports lower subjective psychological well-being measured using a screening tool for depression in December 2020 compared to May and September 2020 (106). Also in keeping with our findings, a representative survey of the German resident adult population showed that a far larger percentage of the population found the overall situation “depressing” in the second lockdown than in the first (39).

A significantly higher proportion of positive PHQ-2 screens (37) and mean symptoms scores (51) in early 2021 compared to 2019 were also found in the German representative panel study (the “Socio-Economic Panel”). However, in contrast to our finding that symptoms of depression first increased in the second wave following an initial decline in the first pandemic months, scores and percentages were actually found to be lower in January/February 2021 than in April through June 2020 in the SOEP. Despite this discrepancy, the January/February 2021 proportion of positive screens in this other study is 12 %, very similar to the September-December 2020 levels (12.1 %) in our study (we do not have data from January and February 2021).

While we have no data on symptoms of anxiety and SRMH from before the pandemic or in its early stages, our findings of a potential increase in symptoms of anxiety and a clear decrease in SRMH between March 2021 and the end of 2021 are in keeping with the picture of worsening mental health following the onset of the second wave.

These changes occurred in the context of a second wave of infections much larger than the first, followed very quickly by a third wave and a fourth, very severe wave with only short periods of lower infection rates in between. Although vaccinations began at the end of 2020, measures to slow transmission were in place for much of this time, mortality rates were high, and hospitals were reported to have come dangerously close to their limits (20, 21, 23-25). While, again, our results do not allow for conclusions on causal relationships between infection rates, mortality, NPIs, or other pandemic factors and mental health, associations between mean PHQ-4 scores and “pandemic intensity” have been reported in a meta-analysis (107).

The sheer increased duration of the cumulative pandemic stressors may also explain potential pandemic-related changes in mental health later on (19). A resilient response is more likely in the face of brief stressors than in the face of more chronic stress (18). In general, longer-term experiences of lack of control and helplessness threaten mental health and may be particularly related to depressive symptoms (16). Reductions in protective factors such as social contact, leisure activities (19), and access to the full spectrum of health services (16) may also grow more harmful with longer durations. Finally, most mental disorders take time to develop and manifest with a prodromal phase (108, 109). While our study does not address the prevalence of any mental disorders, the individual symptoms that comprise these disorders might be subject to the same dynamics.

Our findings of a further increase in symptoms of depression and symptoms of anxiety between late 2021 and early 2022 resulting in by far the highest levels in our over three-year observation period lend further support to the assumption of a potential build-up of pressure on mental health. The pandemic context at this time was a fifth wave of infections driven by the omicron variant immediately following the fourth wave and reaching the largest ever peak in new infections in April 2022 (21, 22), and yet a suspension of most NPIs from the beginning of April (27). Another major and acute population-wide stressor in the final four months of our observation period was the war in the Ukraine beginning on February 24^th^, 2022. It is of note that symptoms of depression and anxiety increased rather markedly in the January/February-centered estimate of 2022, which includes data until mid-March 2022, suggesting potential mental health impacts of the war (110) and emerging economic developments (11, 18). The fact that subsequent estimates remained elevated raises the possibility of developments beyond a short-lived reaction to a discrete event.

#### 4.3.2 Declines in mental health from the second wave onward by subgroup

Turning to subgroups, stratification by sex, age, and level of education in our uniquely long and continuous time series revealed increases in depressive symptom levels at different times after the onset of the second wave of infections in all groups. Gender differences in depressive symptoms did not change over time, but increases in symptom levels were more pronounced in women than in men until the end of 2021. This finding is as expected based on previous literature (e.g. 33, 44, 45, 99) and considering factors such as a greater burden from increased care work among women (45) and increases in domestic violence (14, 16, 45). In 2022, however, men also showed a significant increase above 2019 levels, as well as above 2020 and 2021 levels. This later increase may be due to new stressors or simply a delay in negative mental health developments.

While the sexes and the level of education groups all showed relatively similar overall trajectories, age groups differ in the shape of their time series after the onset of the second wave, suggesting that stressors and protective factors may differ by age in particular. In keeping with previous findings of mental health vulnerabilities among young adults in the pandemic (33, 41, 46-49), the youngest age group stands out in our study for its steep increase in depressive symptoms at the end of 2021, reaching 18.6 % positive screens (standardized estimate, not population estimate) and the highest mean scores among any group at any time. This could be related to the transitional nature of young adulthood, the particularly great importance of social contact with peers when leaving the parental home (19, 43), and an overall greater disruption of life in this group (50) during the pandemic. On the other end of the age spectrum, those aged 65+ years stand out for early significant increases beyond pre-pandemic levels and a particularly constant trend of increase throughout the observation period from 9 % positive screens in spring/summer 2019 to 18 % in 2022 (standardized estimates). While most studies highlight risks among younger adults, a German study using primary care data found early increases of mental health diagnoses among those aged 80 and over (111), consistent with our results. A greater risk of severe disease and death from COVID-19 (16) may have resulted in greater stress and isolation throughout the pandemic (19) in this age group, with less relief from temporary suspensions of NPIs.

The feared widening of disparities in mental health (16, 19, 42) by SES in the pandemic was not evident in our study, which looks at educational differences as one of the dimensions of SES. We found a similar overall trend in all education level groups, with an increase of about five percentage points in each group between spring/summer 2019 and spring/summer 2022. The high level of education group stands out for the greatest relative increase given baseline levels and in terms of how early and continuous increases are across the observation period. Previous international studies from other OECD countries have also found greater increases in psychological distress in higher SES groups (44, 48, 61, 62). In Germany, higher income individuals have, for example, been reported as showing greater declines in life satisfaction during the pandemic (49, 112). Discussed reasons include more working from home in this group (113), which has been shown to be linked to mental health declines in the pandemic (38). Moreover, this group may have experienced a more substantial change in lifestyle more generally (48), perhaps with concomitant greater expectations for the constant availability of resources (62). However, a complex set of risk factors is likely to be at play in all education groups. Occupational and financial difficulties were identified as particularly crucial for an increase in depressive and anxiety symptoms in Germany (38). Importantly, the established social gradient in the risk of depressive symptoms remains unchanged until the end of the observation period in our study, with twice the percentage of positive screens in the low as in the high level of education group in spring/summer 2022.

### 4.4 Strengths and limitations

#### 4.4.1 Strengths

Three features of the study should be highlighted as strengths: **1) Continuous, representative data spanning one year before the outbreak of COVID-19 and over two years of the pandemic:** While most of the existing literature on mental health developments during the COVID-19 pandemic in Germany covers limited time periods and focuses on the early phases of the pandemic only, we present results on the whole course of the pandemic until June 2022, including pre-pandemic data for depressive symptoms. **2) Development of a method for assessing trends at higher temporal resolution:** A method for deriving robust month-by-month results from relatively small samples was developed. Using graphical representations of monthly moving estimates, multiple adjustments of the sample, and smoothing spline curves, we were able to produce graphic time series for the visual identification of trends which were nearly all verified by statistical time period comparisons. This demonstrates the feasibility of this approach to high-frequency mental health surveillance. **3) Examining developments over time both in mean scores and using scale cutoffs**: The relevance of population means for public mental health in connection to Geoffrey Rose’s ideas about prevention and health promotion at the population level has been previously discussed (76). Changes in the population symptom level are of interest irrespective of whether they result in more positive screens. The additional examination of positive screens permits conclusions on whether changes manifest in increases or reductions in cases of potential immediate clinical significance.

#### 4.4.2 Limitations

Limitations in the interpretation and evaluation of our findings include: **1) Time periods of observation and comparison:** Because the time series on anxiety symptoms and SRMH span only about 16 calendar months during the pandemic and include no pre-pandemic data, observed developments cannot be contextualized temporarily and are more difficult to interpret than the longer time series for depressive symptoms (11 months pre-pandemic, 27 months during the pandemic). However, even for depressive symptoms, 11 months of pre-pandemic data are not sufficient to control for seasonal trends and long-term secular trends. **2) Gaps in data collection**: Data collection was interrupted three times for depressive symptoms and once for anxiety symptoms and SRMH. This resulted in four months without data on depressive symptoms and one month on anxiety symptoms and SRMH. These gaps were minimized by using months ranging from the middle of one calendar month to the following calendar month. Additionally, predictions on a three-month window were still made when only two months were included. Thus, results assigned to the central months might be biased towards the first or the third months in the window or just averages of the first and the third month. Also, one gap in the GEDA study was filled with data from the COVIMO study, which had a comparable design but a different overall framing and focus. However, we checked for and did not find systematic, study-related differences. **3) Representativity of the sample for the general population and statistical power for subgroups**: The response to population-based telephone surveys typically varies systematically by sociodemographic factors (114). In particular, younger individuals and those with lower levels of education are underrepresented in our study. We used weighting factors to account for population structure, but the small number of cases in these groups may mean that possible changes over time within a group and differences from other groups might not be detected. In order to reliably achieve statistical significance in the subgroup analyses, larger sample sizes within certain subgroups would be required. Mentally ill and especially severely mentally ill individuals may also be less likely to participate (115, 116), a bias for which we cannot correct. Similarly, we cannot rule out the possibility that willingness to participate in a survey conducted by a governmental public health institute during the pandemic was related to subjective pandemic-related psychological distress. **4) Measurement and scaling**: Using short forms to measure depressive and anxiety symptoms results in a restricted range of scores compared to the full questionnaires. This might have decreased the likelihood of detecting changes compared to the respective long versions with more items. Additionally, the PHQ-2 and the GAD-2 as well as its long versions PHQ-8, PHQ-9, and GAD-7 measure the severity of depressive and anxiety symptoms on an ordinal scale. However, validation supports the interpretation as a metric scale (78, 117). According to this assumption, distributions of PHQ-based measures are commonly described by means of sum scores (e.g. 38, 49, 65, 104). Furthermore, information based on self-report can be subject to recall-bias and social desirability (118).

#### 4.4.3 Conclusion and implications

The **main implications** of our findings derive from the observation of a two-stage substantial decline in mental health in later phases of the pandemic. Importantly, the increase in depressive symptoms in the general population cannot solely be attributed to elevated symptom levels below the clinical screening threshold. Instead, it results in an increase in the proportion of the population screening positive for possible depressive disorder by approximately five to six percentage points when comparing estimates for CW 11-37 in 2019 with almost the same weeks (CW 11-24) in 2022. Our findings of increasing symptoms of anxiety and decreasing SRMH between 2021 and 2022 are consistent with this picture of a deterioration of mental health in the population. Continued surveillance will show whether this deterioration was temporary or part of a more sustained development. The observed developments call for vigilance with regard to possible changes in mental healthcare needs ranging from an increased need for diagnostic clarification and sub-clinical prevention measures to a greater need for secondary prevention. In addition, they point to a great need for mental health promotion and health in all policies approaches (119).

Evidence on **vulnerable groups** can provide guidance in the allocation of measures of mental health promotion and prevention. Overall, none of the examined sociodemographic groups prove to be consistently resilient. An effective public health response thus faces the challenge of addressing the entire population and cannot target clearly identifiable risk groups. However, in keeping with many other studies, a particular focus on women and young adults, but also the eldest adults, may be warranted. The final months in our time series, which saw the introduction of a new major societal-level stressor, indicates that mental health developments of men and adults in later middle age should also be observed closely. They show that vulnerabilities may be subject to change over time, demanding continued observation and reporting to increase awareness and flexibility in public health policy and mental health practitioners. As was the case before the pandemic, there is still a high need for mental health support for individuals of low socioeconomic status. Despite our finding of particularly early and continued increases in depressive symptoms among the high level of education group, the social gradient of lower mental health in the low level of education groups clearly persists across our time series.

##### Particularly with regard to current circumstances, mental health trends in the population should be observed and evaluated continuously and systematically

Further temporal dynamics in mental health seem very likely in view of a wide range of potential contributing factors and ongoing crises. These include the continued dynamic development of the pandemic and public health measures in response (120), the risk of chronification of stress reactions due to the persistence of stressors or loss of resources (16, 19), and the emergence of further mental health risk factors such as a long-term economic recession (11, 18) as well as other crises not related to the pandemic. It is possible that recent events such as the war in the Ukraine may have contributed to 2022 mental health declines (110). The exacerbation of the global climate crisis (121, 122) represents another major ongoing contextual factor. All of these crises taken together might also contribute to increased experiences of multiple disasters, which can exert a specific impact on public health (123).Fundamentally, the psychological impact of crises is likely to vary over time. For the pandemic, we can assume overlapping effects of immediate fear, followed by responses to adversities, consequences of insufficient mental health support, and long-term implications of recession or uncertainty (100). Because mental disorders frequently develop over a longer period of time during which multiple stressors exceed individual resources and interact with individual vulnerability, the possibility of delayed and substantial rises in the prevalence of mental disorders cannot be ruled out.

##### A continuation of mental health surveillance made possible by uninterrupted data collection is also needed in less tumultuous times to safeguard crisis preparedness

Our results show that mental health trends in the general population can change suddenly, supporting the utility of an early warning system. Sufficiently long time series of mental health indicators are required in order for high-frequency surveillance to help inform public health policy by identifying changes, assessing their significance and relevance against the backdrop of previous dynamics, and evaluating the impact of public health interventions effectively. In addition to this fundamental need for continuous mental health data, **future studies should** expand findings to the whole life span by including the observation of children and adolescents. Moreover, they should go beyond the use of screening instruments to measure symptoms in assessing the prevalence of mental disorders and include longitudinal designs in order to better understand mechanisms of vulnerability and resilience in the face of individual as well as collective determinants of mental health.

## Data Availability

Population-based data from the German health monitoring program that support the findings of this study are available from the Robert Koch Institute (RKI) but restrictions apply to the availability of these data, which were used under license for the current study and so are not publicly available. The data set cannot be made publicly available because informed consent from study participants did not cover public deposition of data. However, a minimal data set is archived in the Health Monitoring Research Data Centre at the RKI and can be accessed by all interested researchers. On-site access to the data set is possible at the Secure Data Centre of the RKIs Health Monitoring Research Data Centre. Requests should be submitted to the Health Monitoring Research Data Centre, Robert Koch Institute, Berlin, Germany.

## 5 List of abbreviations

COVIMO: COVID-19 vaccination rate monitoring in Germany
CW: calendar week
GAD-2: Generalized Anxiety Disorder Questionnaire (2 items)
GEDA: German Health Update
EHIS: European Health Interview Survey
PHQ-2/8/9: Patient Health Questionnaire (2/8/9 items)
RKI: Robert Koch Institute
SES: Socio-economic status
SRMH: Self-rated mental health

## 6 Declarations

### 6.1 Ethics approval and consent to participate

GEDA and COVIMO are subject to strict compliance with the data protection provisions set out in the EU General Data Protection Regulation (GDPR) and the Federal Data Protection Act (BDSG). Participation in the study was voluntary. The participants were informed about the aims and contents of the study and about data protection. Informed consent was obtained verbally. In the case of GEDA 2019/2020, the Ethics Committee of the Charité –Universitätsmedizin Berlin assessed the ethics of the study and approved the implementation of the study (application number EA2/070/19).

### 6.2 Consent for publication

Not applicable

### 6.3 Availability of data and materials

Population-based data from the German health monitoring program that support the findings of this study are available from the Robert Koch Institute (RKI) but restrictions apply to the availability of these data, which were used under license for the current study and so are not publicly available. The data set cannot be made publicly available because informed consent from study participants did not cover public deposition of data. However, a minimal data set is archived in the Health Monitoring Research Data Centre at the RKI and can be accessed by all interested researchers. On-site access to the data set is possible at the Secure Data Centre of the RKI’s Health Monitoring Research Data Centre. Requests should be submitted to the Health Monitoring Research Data Centre, Robert Koch Institute, Berlin, Germany.

### 6.5 Author Contributions

EM and LW contributed equally. EM and LW wrote most sections of the manuscript, integrating drafts for specific sections from co-authors. EM developed the main conceptual ideas and coordinated the research process as a senior researcher. EM, LW, SJ, SD, CK, and SM developed the specific research questions and discussed the analytical approach. SJ performed the statistical analyses, worked with SD to develop the statistical methods, and produced the results graphs.Together they drafted the analysis section of the manuscript. LW and EM interpreted the results. CK primarily contributed to the interpretation of educational disparities, assisted by SM as senior researcher in the area of health inequalities. JT, as project leader of the Mental Health Surveillance, contributed to all content discussions and by drafting some sections of the Introduction and Discussion. SE contributed information on the existing relevant literature for Germany. DP and SuS contributed to the manuscript with comments and suggestions, as did HH as the head of the Unit of Mental Health at the RKI. All authors gave crucial input on drafts of the manuscript and read and approved the final version.

## 6.7 Acknowledgements

We would like to thank all colleagues in Unit 21: Epidemiological Data Center of the Department RKI who provide us with quality-assured data on a monthly basis. In particular, Jennifer Allen as project manager of the GEDA surveys has collaborated continuously.

